# Households at Higher Risk of Losing at Least One Individual in India: if COVID-19 is a new normal

**DOI:** 10.1101/2020.06.08.20125203

**Authors:** Rajeev Ranjan Singh, Palak Sharma, Priya Maurya

## Abstract

After the outbreak of COVID-19 and the passing of a few months with this pandemic; the world has started to adopt strategies to live with the virus. The WHO has also accepted that the pandemic caused by the novel coronavirus is going to last longer, and suggested that one needs to learn to live with this virus. Accepting this bitter truth that this pandemic is going to be a new normal and people of all ages can be infected by the new coronavirus; however, older people and those with chronic diseases are more vulnerable to the virus. The study tries to access the household with at least one patient with few selected chronic diseases in India, which are presumed to be at a higher risk of losing at least one individual if this pandemic scenario is going to last long and spread is wider. The study used nationally representative data (NSSO) for information on morbidity and other health-related issues. Data from the official website of the Ministry of Health and Family Welfare dedicated to COVID-19 reports have been used to look into the recent happenings caused by COVID-19 pandemic in India. Bivariate analysis has been used to calculate household at risk, and binary logistic regression has been used for the likelihood of household at risk. The case-fatality ratio is calculated using the number of confirmed cases and the number deceased due to the same. The study found that about 9.4% of Indian households are at a higher risk of losing at least one individual. Older people (60+), males and households with better economic status are at a higher risk. The chronic condition varies by states and social-economic and demographic status. The share of households at higher risk was highest in Kerala (33.19%), followed by Andhra Pradesh (19.85%) and Chandigarh (19.05%).

## Introduction

The outbreak of the rapidly spreading severe acute respiratory syndrome coronavirus 2 (SARS-CoV-2), also known as COVID-19, was first reported in Wuhan city of China in December 2019. Since then, it has escalated all over the world within a very short span of time and has literally brought a significant challenge to the public health system globally with no exception in India. With a very high infectivity rate and rapid increase in the deceased cases, the virus has impacted a significant chunk of the population worldwide. Globally as of 21^st^ May 2020; about 4904413 people have been affected with COVID-19, and around 323412 have lost their lives due to the pandemic (https://covid19.who.int). In India, the very first case was reported on 30^th^ January in the state of Kerala, and on the same date, the World Health Organization (WHO) Emergency Committee declared a global health emergency based on growing case notification rates in China and other international locations. Later on 11^th^ March 2020, the WHO characterized novel coronavirus as a global pandemic. In India, as on 22^nd^ May 2020, the number of confirmed cases of COVID-19 reached a tally of 118447 patients and is one among 11 countries to cross 1 lakh confirmed cases. A total of 3583 patients weren’t lucky enough to survive it. There has been an apparent disparity in the number of cases and their spread within the states of India. As per WHO’s latest situation report number 125, India hasn’t reached to the community transmission mode and still has “clusters of cases”.

With this very unpredictable scenario of COVID-19, nations are still trying to absorb about the pandemic. According to the WHO’s guidelines people of all ages can be infected by the new coronavirus (COVID-19), however, the risk is higher for a particular section of the population. Recent studies carried out on the characteristics of patients (using hospital data) and using meta-analysis have shown that most common non-communicable diseases associated with COVID-19 disease were hypertension, diabetes and cardiovascular diseases (Du Y et al., 2020; Lupia T et al, 2020; Fauci et. al, 2020) and these were significantly higher among those who were severely ill/died of COVID-19 as compared to not so critically ill (Zheng Z et al., 2020; Singh AK et al., 2020). Other comorbidities that are highly associated with COVID-19 are cancers, strokes and, respiratory infections including asthma and chronic obstructive pulmonary disease (COPD) (Singh AK et al., 2020; Zheng Z et al., 2020; Clark A et al., 2020; Kunal S et al., 2020; Lake M. A. 2020; Kiaghadi, A et al., 2020; Yao Q et al. 2020). According to global estimates, most of the mortality due to COVID-19 attributed to either one or more comorbidities (Fang et al., 2020; Leung et al., 2020).

As per GBD 2017 India estimates, NCDs have the largest share in deaths (63%) and around every three out of four people (74%) aged 50 or above die to due to one or the other non-communicable disease. The challenge of tackling parallelly new COVID-19 cases and maintaining continuity of healthcare for NCD patients in the time of pandemic knowing that people suffering with NCDs are at higher risk is a gigantic task for the public health sector. As per reports from the Ministry of Health and family welfare, among the total patients deceased, 70% had comorbidities. Since there are certain sections of the population that are at higher risk of getting affected than the others and it is important to identify people who may be at most risk of COVID-19 to inform policy and intervention.

However, WHO has accepted that the pandemic caused by the virus is going to last longer, and has suggested that one need to learn to live with this virus. This study aims to identify the proportion of households at a higher risk of getting affected by COVID-19 given this pandemic continues to affect the nation at today’s pace. Accepting this bitter truth that this pandemic is going to be a new normal and people with chronic disease are more vulnerable to the virus. This study also examines the factors that affect the households for being at risk of getting affected by one or more comorbidities.

## Data and Methods

The study used a national representative data conducted by National sample survey organisation (NSSO), Ministry of Statistics and Programme Implementation on household social consumption related to Health during the period July 2017 to June 2018, namely schedule 25.0 of 75^th^ round survey. The main objective of the survey was to gather basic quantitative information on morbidity, profile of ailments including other health related issues. The survey covered nationwide 113823 households where 89410 households have at least one patient (including inpatient and outpatient). (NSSO, 2018). We also used data from the latest reports on COVID-19 from the official website of Ministry of Health and Family Welfare (https://www.mohfw.gov.in/). The study has classified households in two categories and defined “households at risk” as a dependent variable. It takes value ‘1’ if at least one patient in the particular HH is suffering from chronic disease (Tuberculosis, cancer, diabetes, hypertension, heart disease, and respiratory diseases) and takes the value ‘0’ if no-one in the household have any patient with chronic diseases. There are various socio-economic and demographic factors that affect households being affected by chronic illness like the age of patient, gender, the place where he/she resides, wealth quintile, level of education, size of the household, religion and region one belongs from.

### Statistical Analysis

The proportion of households at risk by different social-economic characteristics is presented using bivariate analysis. The bivariate logistic regression model is fit predicting the likelihood of household at risk. Binary logistic is used when our dependent variable is categorical with binary response (1 “household at risk, 0 “Household without risk”). The case fatality rate (CFR) is calculated which is a measure of the severity of a disease and is defined as the proportion of deaths due to a COVID-19 per 100 confirmed cases.

## Results

Out of a total surveyed households (113823) about 9.38% (N=12891) of the households are found to be at risk where atleast one person in the household is suffering from one or more of the chronic diseases. **Figure 1** shows the percentage distribution of chronic diseases in surveyed households. Among all the reported chronic ailments the highest proportion was shared by hypertension (28%) followed by diabetes (27%) and respiratory diseases (25%) and least shared by tuberculosis (2.24%).

**Figure 1:**
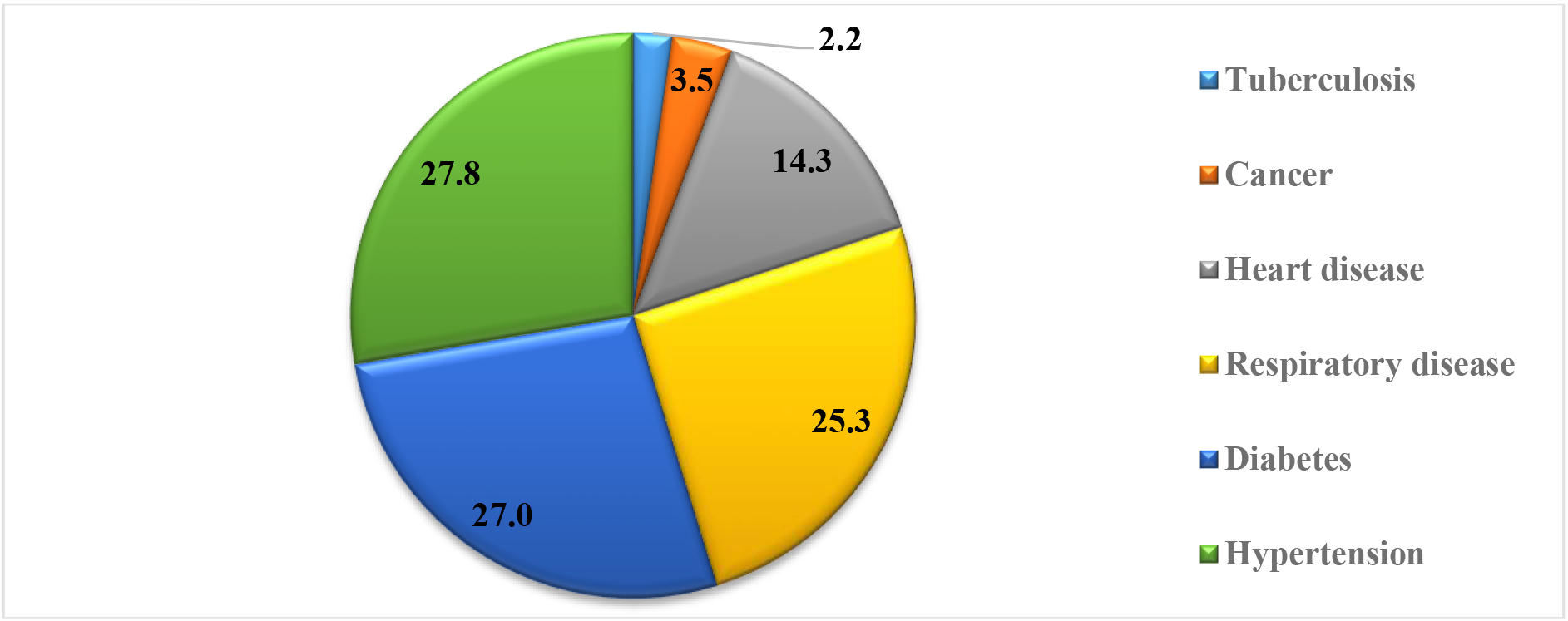
Percentage distribution of chronic diseases in the surveyed households, India, 2018.

**Table 1** provides the descriptive statistics of households with higher risk of major morbidity in India. Results show that population aged 60 and above are at the highest risk of having one or more chronic diseases (52.3%). Every two out of five (40%) belonging to age group 45-59 have at least one morbidity condition. The risk is predominantly higher in men (33 percent) as compared to their female counterpart (20.5%). The urban population is at higher risk with about 12% of households being at risk while in the rural area; about 8 percent of households have people with one or more chronic comorbidity. As the wealth quintile increases; the proportion of households at risk (with at least one member having chronic illness) increases. One-fourth household those had a better economic status (including rich and richest wealth quintile), are at higher risk. Education of the household head doesn’t show a particular pattern where and shows that household with no education (28%) and higher education (27%); both are at higher risk of being in one or more chronic morbid state. With an increase in the household size, the risk of atleast one person having one or more comorbidities is higher. Household with 1-4 individuals are at lower risk (9.2%) as compared to the households with more than 9 members (10.8), although the gap is narrow. The results reveal that about 11% of the households that follow Muslim religion are at a higher risk as compared to those who follow Hindu religion (9%). The highest risk of a HH being at risk is in the southern region (13%) followed by eastern and western regions (9% each). A notable small percentage of the household from the north-eastern region (2.9%) are at higher risk.

**Table 1:** Percent of household at risk of chronic diseases with background characteristics of the households, India, 2018 (N=12891)

| **Background Characterstics** | **Percent of HH at Risk** |
| --- | --- |
| **Age** |  |
| <15 | 13.97 |
| 15-44 | 10.37 |
| 45-59 | 40.82 |
| 60+ | 52.25 |
| **Gender** |  |
| Men | 32.95 |
| Women | 20.50 |
| **Type residence** |  |
| Rural | 8.01 |
| Urban | 12.21 |
| **Wealth Quintile** |  |
| Poorest | 6.07 |
| Poorer | 8.12 |
| Middle | 8.25 |
| Richer | 9.95 |
| Richest | 12.98 |
| **Level of education** |  |
| Illiterate | 28.19 |
| Up to Primary | 24.8 |
| Middle/Secondary | 22.79 |
| Higher secondary & above | 27.48 |
| **Size of Household** |  |
| 1-4 | 9.21 |
| 5-8 | 9.50 |
| 9+ | 10.81 |
| **Religion** |  |
| Hindu | 8.91 |
| Muslim | 10.86 |
| others | 13.59 |
| **Region** |  |
| Northern region | 7.73 |
| Eastern region | 8.83 |
| Northeast | 2.92 |
| Western Central | 8.9 |
| Southern region | 12.76 |
| **Total** | 9.38 |

**Table 2** represents the results of the binary logistic regression. The results show that higher age significantly related to higher risk. The odds of household being at risk was around 8.6 times (95% CI: 8.554-8.580) higher for respondents in age-group 60+ and 5.4 (95% CI: 5.440-5.455) times higher for those in age-group 45-59 compared to reference category (15-44 years). Risk for men is 34% higher (95% CI: 1.348-1.351) as compared to their female counterparts. Compared to rural residents, the odds of households in urban areas being at higher risk is 1.61 times greater (95% CI: 1.609-1.613). Compared with the household belonging to a more deprived class, the odds of a household being at a higher risk of having one or more comorbidities significantly increases with a better economic situation. Household with the richest wealth quintile was 1.72 times (95% CI: 1.723-1.729) more likely to be at risk. As compared to illiterate population; people with higher education have the highest probability of reporting at risk (OR: 1.17; 95% CI: 1.168-1.172). Households that belong to Muslim religion are 35% (95% CI: 1.354-1.358) more likely to be at higher risk for disease compared to their Hindu counterparts. As far as the regions are concerned; households from the southern region are 2.08 times (95% CI: 2.079-2.097) more likely to be at a higher risk from the disease compared to the household from the north-eastern region. Household size was positively associated with higher risk from the disease.

**Table 2:**
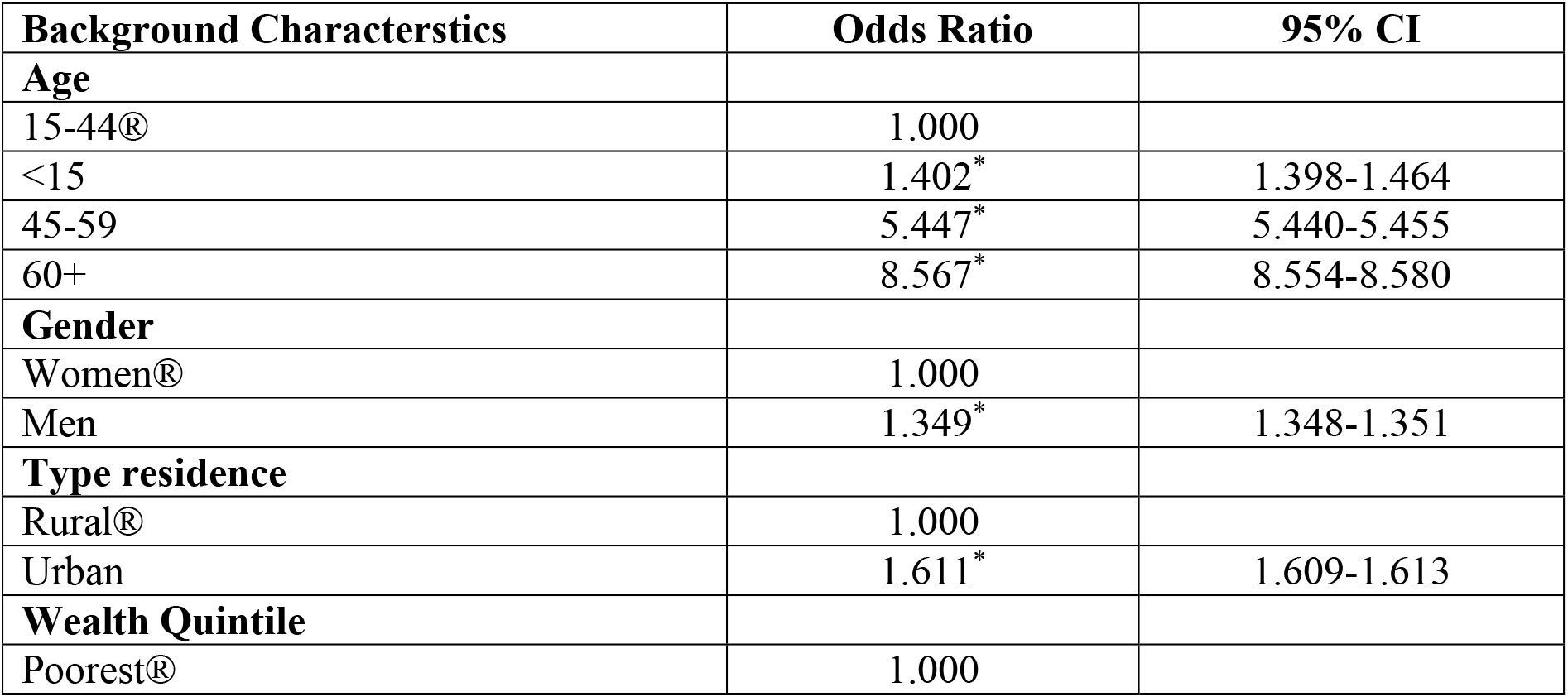
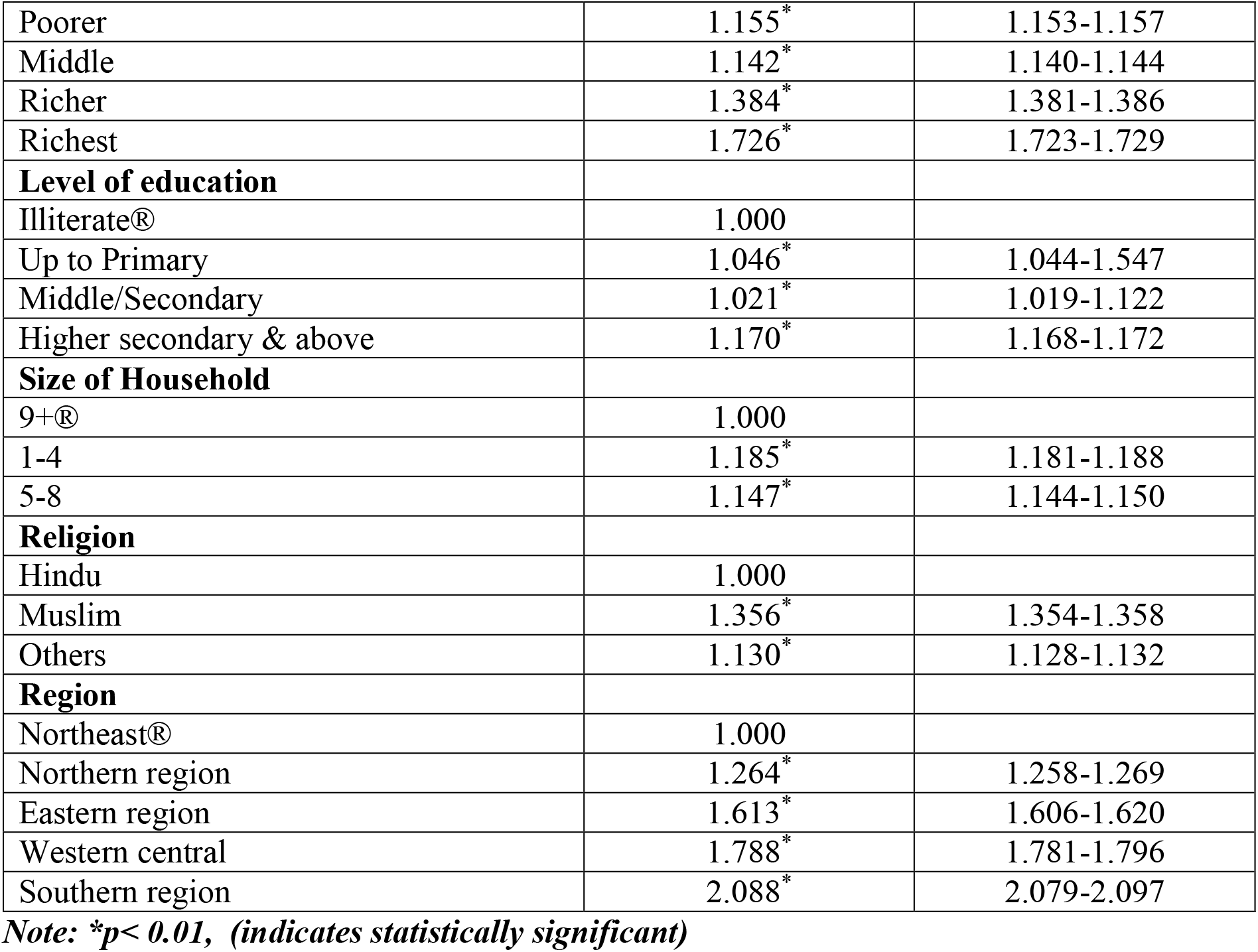
Odds ratio of household at higher risk with background characteristics, India, 2018.

**Figure 2** shows the proportion of households at higher risk of one or more chronic morbidity in different parts of the country. Households with higher risk are not evenly distributed. Mainly southern part of India had household with higher risk of morbidity with Kerala at the top of the list with 33% of the households at higher risk. Other states that have higher proportion of households at risk are Andhra Pradesh (19.82%), Goa (15.89%), West Bengal (16.69%) and Punjab (15.51%). The prevalence of the household at higher risk was comparatively lower in north-eastern parts of the country. **Figure 3** shows that number of confirmed cases of COVID-19 as of 22 may,2020. Highest number of cases of COVID-19 are reported in the state of Maharashtra where the number has reached to 41642. Tamil Nadu and Gujarat are also at the top with more than 10000 positive cases of COVID 19. Central and western part of the country are mainly affected and have reported with higher number of cases so far other than any part of the nation, while north-eastern region reported lowest cases. **Figure 4** shows the case fatality rate per 100 confirmed patients in the different states in India. The figure shows that fatality rate is highest in West Bengal followed by Maharashtra and Gujarat. (for detailed numbers and % for figures 2-4 see **Appendix A1**).

**Figure 2:**
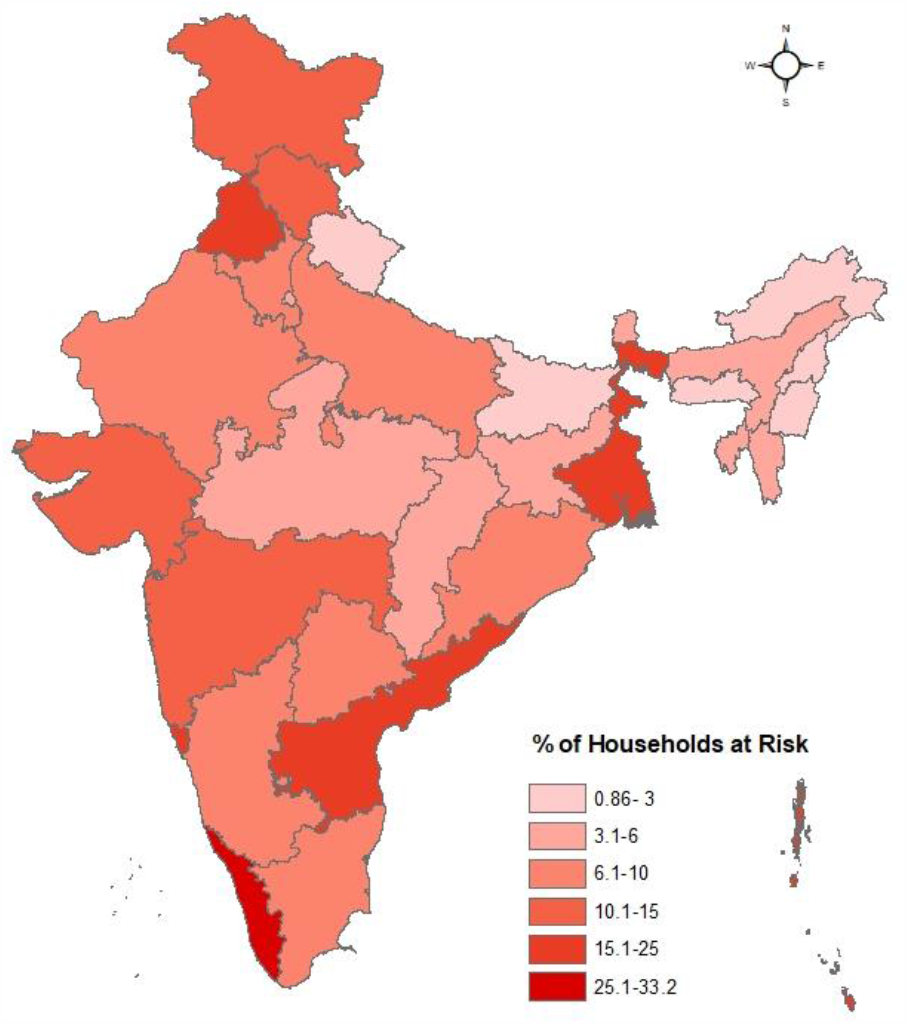
Proportion of households at risk across states of India, 2018.

**Figure 3:**
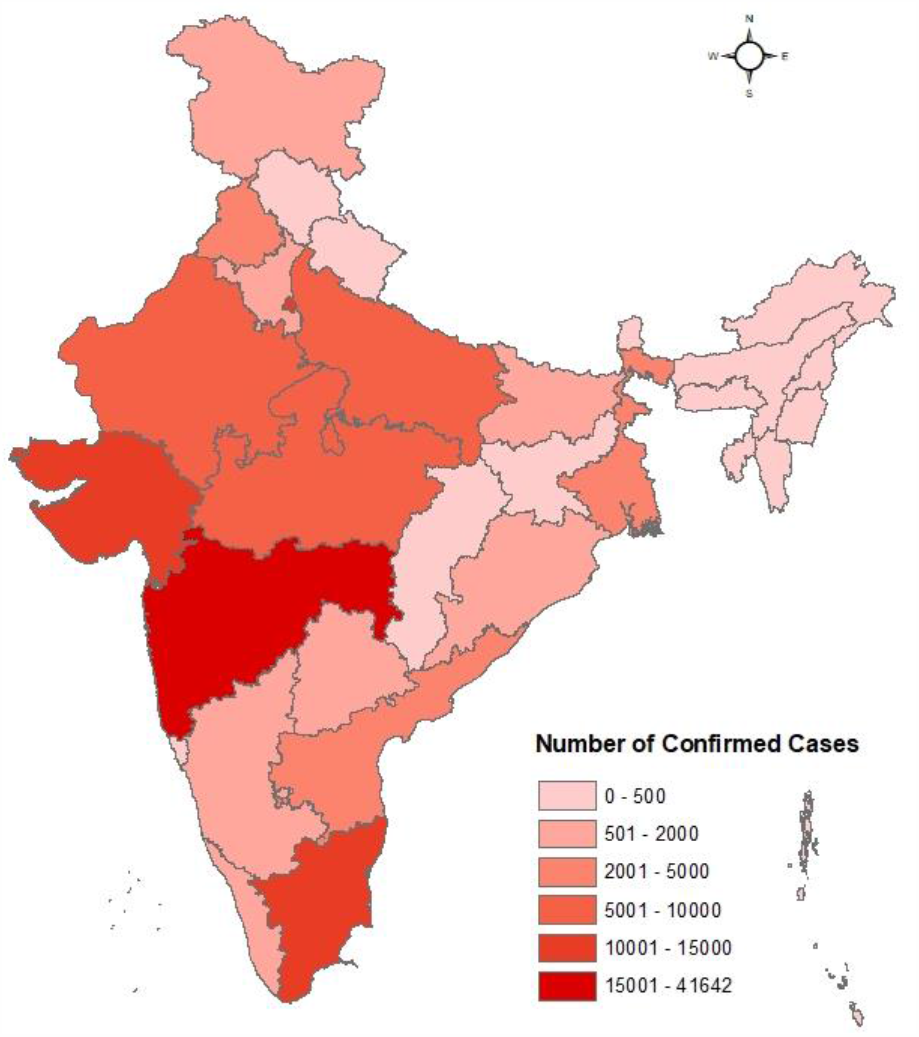
The total number of confirmed positive cases of COVID-19 in India, 2020.

**Figure 4:**
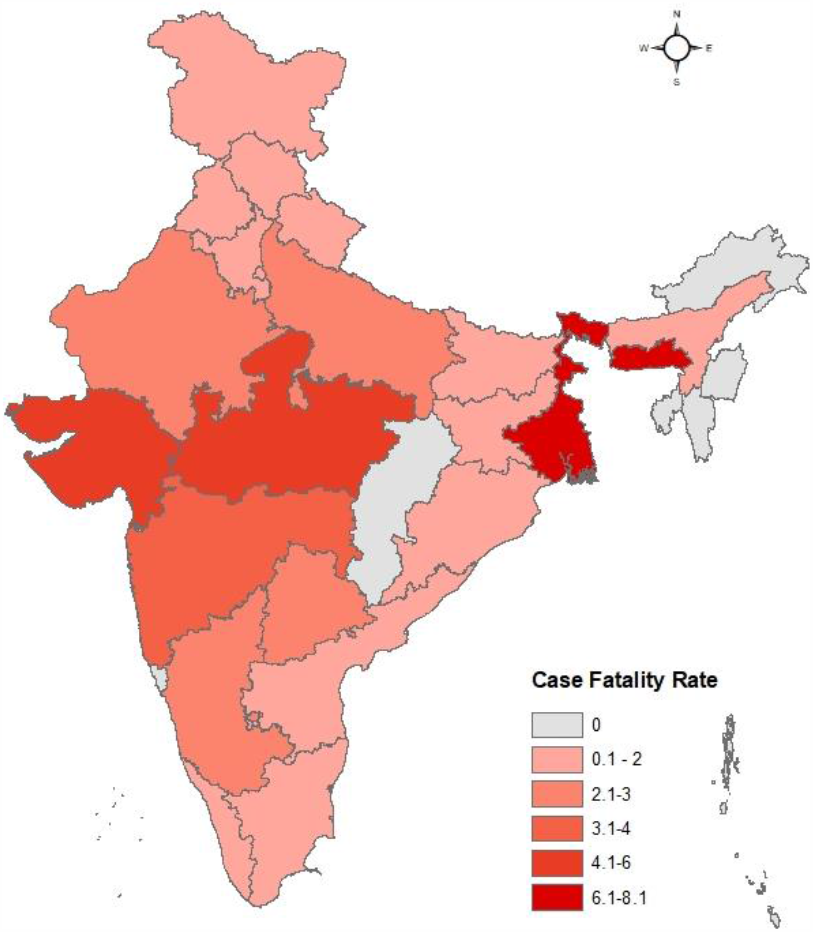
The case fatality rate per 100 confirmed cases of COVID-19 in the states of India, 2020.

## Discussion and Conclusion

The rapidly increasing cases of COVID-19 all over the world and upsurge in the deceased patients have literally challenged the public health system of all the nations. The WHO has accepted that the pandemic caused by the novel coronavirus is going to stay long and one needs to learn to live with this virus. It is being revealed that however anyone can get affected by the coronavirus but those who are old (60+) and those having pre-existing chronic disease are at a higher risk. Given that it is important to identify the proportion of population that may be at most risk of COVID-19 to inform policy and intervention. The study has identified the proportion of households that are at higher risk of getting affected by COVID-19 and may loose atleast one family member given that atleast one of the household member is suffering from one or more chronic comorbidity.

Overall about 9.38% of all the survey households are at a higher risk of loosing atleast one member of the family who is suffering from one or more comorbidities like diabetes, hypertension, respiratory infections and heart disease. A study done by Guan et. al (2020) shows that 25 percent COVID-19 patients reported having at least more than one comorbidity, and the most prevalent comorbidity was hypertension followed by diabetes. Previous study from different countries shows that older age people, men, having hypertension, diabetes, heart failure and cardiovascular disease are more prone to COVID-19 disease [Shi et al., 2020; Bonow et al., 2020; Grasseli et al., 2020; Zeng Z et al., 2020; Yao Q et al., 2020]. The median age for the disease is 51 years and this is higher for male (52 years) and lower for females (50 years) (WHO report, 2020).

Countries with a high old-age dependency ratio show higher fatality rates due to this disease. COVID-19 severity is higher in those countries where there is already an aging population, with higher risk and requirement of hospilization [Sen, K., 2020]. In New York, 10 percent of total population and 16 percent of senior population are vulnerable and at highest risk of COVID-19 [Kiaghad et al., 2020] and similar results from present study indicates that the household with older people and males are at higher risk of developing a chronic morbidity and hence are at increased risk of severe COVID-19 disease. Other studies have also shown that more males have been reported to have COVID-19 and majority among the deceased (73%: Lake M 2020, Zheng Zet al., 2020). Household in urban area are at higher risk as the odds of a person having chronic morbidity is higher in urban areas as compared to rural settlement. Although a study by Corburn et al., 2020 reported that households of informal settlements like slum, vulnerable urban agglomeration, are least prepared for this pandemic since basic WASH facilities (water, sanitation, hygiene) adequate household and public space are not available properly.

The finding of the study shows that socioeconomically disadvantaged households at lower risk for comorbidities compare to households with economically better off, but in this scenario, socioeconomically backward class are most vulnerable for failing to meet healthcare and WASH facilities [Basu S, 2020]. There is a wide regional variation in the burden of COVID-19 cases. The risk is higher in the states of Kerala, West Bengal, Punjab and Andhra Pradesh given they have higher proportion of households with pre-existing comorbidities. Kerala being a developed state in terms of healthcare has comparatively less number of cases of COVID 19 and has fever fatality. On the other hand, states like Maharashtra, Delhi, and Gujarat are still trying to cope up and have highest number of confirmed cases. State of West Bengal being one of the lower HDI states and has high population density; has high number of households at risk and also shows high case fatality rate. It is observed that in high HDI states like Kerala, Chandigarh and Punjab, the growth of disease is slow and flattering the curve while disease is increasing in low and mid HDI states. This might be the cause of public healthcare facilities [Gupta R et al., 2020].

With every phase of lockdown in India there come new strategies and a few relaxations by the central or state government. This study will help the policymakers to identify the states/districts where the risk is potentially higher for becoming the next hot spots given where the household with risk are highest. This will help to mitigate the impact of COVID-19 on the high-risk population by preparing for future situations of the pandemic. With the given heightened risk due to coronavirus; special care is required to patients who are vulnerable (with associated comorbidities) and are at a higher risk of getting affected. Changes in personal behaviour and norms could be required for months to overcome the hard times.

**Limitation of the study**, we compared the the spread and case fatality ratio of COVID-19 cases happening in the year 2020 with the surveyed population in the year 2018 assuming that there will be no major changes in the socio-economic and demographic patterns of the particular region and states of india in past two years.

## Data Availability

The study used nationally representative data conducted by the National sample survey organization (NSSO), Ministry of Statistics and Programme which is publically available on the official NSSO website.

## Appendix A1: Number of Cases and deaths due to COVID-19 in each of the states of India with Case fatality rate and proportion of households at risk in India

| S. No. | States | Number of Confirmed Cases | Deaths | Case fatality rate | Proportion of Households at risk |
| --- | --- | --- | --- | --- | --- |
| 1 | Andaman and Nicobar Islands | 33 | 0 | 0 | 18.98 |
| 2 | Andhra Pradesh | 2647 | 53 | 2 | 19.82 |
| 3 | Arunachal Pradesh | 1 | 0 | 0 | 2.29 |
| 4 | Assam | 203 | 4 | 1.97 | 3.01 |
| 5 | Bihar | 1982 | 11 | 0.55 | 1.93 |
| 6 | Chandigarh | 217 | 3 | 1.38 | 19.05 |
| 7 | Chhattisgarh | 128 | 0 | 0 | 5.6 |
| 8 | Dadar Nagar Haveli | 1 | 0 | 0 | 0.86 |
| 9 | Daman & Diu | 0 | 0 | - | 6.5 |
| 10 | Delhi | 11659 | 194 | 1.66 | 4.05 |
| 11 | Goa | 52 | 0 | 0 | 15.89 |
| 12 | Gujarat | 12905 | 773 | 5.99 | 13.35 |
| 13 | Haryana | 1031 | 15 | 1.45 | 7.26 |
| 14 | Himachal Pradesh | 152 | 3 | 1.97 | 14.49 |
| 15 | Jammu and Kashmir and Ladakh | 1493 | 20 | 1.34 | 14.82 |
| 16 | Jharkhand | 290 | 3 | 1.03 | 5.55 |
| 17 | Karnataka | 1605 | 41 | 2.55 | 6.43 |
| 18 | Kerala | 690 | 4 | 0.58 | 33.19 |
| 19 | Lakshadweep | 44 | 0 | 0 | 23.1 |
| 20 | Madhya Pradesh | 5981 | 270 | 4.51 | 5.05 |
| 21 | Maharashtra | 41642 | 1454 | 3.49 | 12.7 |
| 22 | Manipur | 25 | 0 | 0 | 2.86 |
| 23 | Meghalaya | 14 | 1 | 7.14 | 0.98 |
| 24 | Mizoram | 1 | 0 | 0 | 5.07 |
| 25 | Nagaland | 0 | 0 | - | 1.59 |
| 26 | Odisha | 1103 | 7 | 0.63 | 8.86 |
| 27 | Puducherry | 20 | 0 | 0 | 4.78 |
| 28 | Punjab | 2028 | 39 | 1.92 | 15.51 |
| 29 | Rajasthan | 6227 | 151 | 2.42 | 6.33 |
| 30 | Sikkim | 0 | 0 | - | 5.59 |
| 31 | Tamil Nadu | 13967 | 94 | 0.67 | 9.92 |
| 32 | Telangana | 1699 | 45 | 2.65 | 6.12 |
| 33 | Tripura | 173 | 0 | 0 | 4.33 |
| 34 | Uttar Pradesh | 5515 | 138 | 2.5 | 7.12 |
| 35 | Uttarakhand | 146 | 1 | 0.68 | 2.91 |
| 36 | West Bengal | 3197 | 259 | 8.1 | 16.69 |

